# Why the SARS-CoV-2 antibody test results may be misleading: insights from a longitudinal analysis of COVID-19

**DOI:** 10.1101/2020.12.08.20245894

**Authors:** Jorg Taubel, Samuel Thomas Cole, Christopher S. Spencer, Anne Freier, Dorothée Camilleri, Ulrike Lorch

**Author notes:** Correspondence to : J Taubel.

## Abstract

To estimate the effectiveness of vaccines in development, a robust mechanism is required to understand immunity, risks of reinfection and measure the immune response to severe acute respiratory syndrome coronavirus 2 (SARS-CoV-2), and how this may change over time. This study is a longitudinal analysis of COVID-19 infection rates using PCR, membrane immunoassay and chemiluminescent microparticle immunoassay (CMIA) diagnostic tests. Our data confirm that antibody levels wane in the three months after symptom onset. Comparison of the three methods used suggests that quantitative CMIA testing may exaggerate numbers of COVID-19 negative individuals.

## Introduction

SARS-CoV-2 emerged in late 2019 as the causative agent for COVID-19. It is an ongoing pandemic shown to cause a range of clinical outcomes – from asymptomatic infection to severe respiratory distress and death. The development of immunity is uncertain and so is the connection with antibody levels. A simple and robust test is important so that current and past infections can be monitored. Having a simple test has become even more urgent now, as we otherwise have no way of monitoring the waning of the vaccine’s efficacy. In cases of an established infection, IgG antibodies are found between 8 and 28 days from onset of symptoms. Their levels then fall over a period of 3-6 months to very low or undetectable levels [1]. However, by using semi-quantitative tests using a fixed cut off, only negative or positive results are obtained.

Numerous longitudinal studies reflecting the state of SARS-CoV-2 infection and immunity have been published [1,2]. These studies suggest an initial acquired immune response in which IgG titres specific to SARS-CoV-2 proliferate over 15 days post-infection, followed by waning immunity to SARS-CoV-2 over the two to three months that follow. The aim of this study was gain a better insight into duration of immunity to SARS-CoV-2 and determine the reliability of a membrane-based qualitative assay to measure past infection. Our study corroborates findings that antibody levels wane over the initial 90 days after infection and provides further insights into how antibody testing for SARS-CoV-2 may underestimate numbers of previously infected individuals.

## Methods

### Participants and testing methodologies

This is a longitudinal analysis on SARS-CoV-2 infection using polymerase chain reaction (PCR) testing to detect active infection, and two different antibody tests – a membrane-based qualitative assay to measure past infection, and a quantitative chemiluminescent microparticle immunoassay (CMIA) to quantify antibody titres. Our study participants fell within two cohorts: staff members, and visitors and trial participants participating in other clinical trials. Cohort demographics are shown in **Table 1**.

**Table 1.** Demographic data for trial participants.

|  | Staff Cohort<br>n = 249 | Visitors and Volunteers<br>n = 1591 |
| --- | --- | --- |
| **Sex, n (%)** |  |  |
| Male | 117 (47.0) | 1038 (65.2) |
| Female | 132 (53.0) | 553 (34.8) |
| **Race, n (%)** |  |  |
| Black African | 20 (8.0) | 90 (5.7) |
| Black Caribbean | 9 (3.6) | 49 (3.1) |
| Caucasian | 147 (59.0) | 1170 (73.5) |
| Chinese | 2 (0.8) | 5 (0.3) |
| Hispanic | 3 (1.2) | 7 (0.4) |
| Indian (India) | 11 (4.4) | 15 (0.9) |
| Japanese | 21 (8.4) | 110 (6.9) |
| Malaysian | 0 | 1 (0.1) |
| Mixed Race | 11 (4.4) | 67 (4.2) |
| Other Asian | 18 (7.2) | 48 (3.0) |
| Other Far Eastern | 2 (0.8) | 11 (0.7) |
| Pakistani | 0 | 1 (0.1) |
| Taiwanese | 4 (1.6) | 17 (1.1) |
| **Age (years)** |  |  |
| Mean (SD) | 34.3 (10.5) | 40.1 (18.0) |
| Median | 31 | 34.0 |
| Range | 20, 66 | 3, 89 |

To gain a better understanding of the duration of immunity, we quantified IgG levels of 21 individuals after they were infected with SARS-CoV-2 using the Abbott Laboratories (Illinois, USA) chemiluminescent microparticle immunoassay (CMIA). The test works by binding to SARS-CoV-2-specific IgG antibodies in a blood sample. Upon binding, the reaction substrate generates luminescence, which is measured by the Abbott Architect system. This luminescence is directly proportional to SARS-CoV-2-specific IgG antibody concentration and is quantified by signal/cut-off (S/CO) units [4]. The Abbott CMIA is technically a qualitative assay, with values above 1.4 S/CO units reported as SARS-CoV-2 positive. This cut-off value has been calculated to maximise positive predictive values (PPV) and minimise false positives, according to the manufacturer. Public Health England found that specificity was 100%, but sensitivity was 93% [5]. In principle, the CMIA test should be able to quantify SARS-CoV-2-specific IgG antibody titres, which would be a more useful use of the system.

### Statistical Analysis

Descriptive statistics were used to summarise quantitative data. Both a linear regression and a local regression (LOESS) fitting methods were used when displaying graphically the IgG values over time after PCR positive test. A log-linear modelling approach was used to calculate the population half-life, accounting for repeated measures.

### Ethical statement

This brief report communicates results taken from a study reviewed and given favourable opinion by NRES Committee (West Midlands - Edgbaston) (IRAS ID: 281788).

## Results

### PCR and Membrane Immunoassay Testing

To date, we have tested a total 1840 individuals (repeat testing, between 1 and 82 times), for a total of 13180 PCR tests, and 6061 antibody tests. Of the 13180 PCR tests conducted, 12641 (96%) were negative, and 91 (1%) were positive. We observed 12 new infections during the week of March 26 2020 - April 1, 2020, our highest observed rate of infection (**Figure 1)**. This was comparable to broader statistics recorded in the UK, where the highest number of daily new cases of COVID-19 in the initial peak was reported on April 12, 2020 (8719 new confirmed cases) [3]. The initial England peak was sustained until the end of April, however the peak we observed quickly subsided and dropped to zero new positive cases by the week 16 April 2020 - 23 April, 2020. Whilst we have observed an increased number of new cases in October, we have not seen the same secondary increase in incidence as seen in the rest of England.

**Figure 1.**
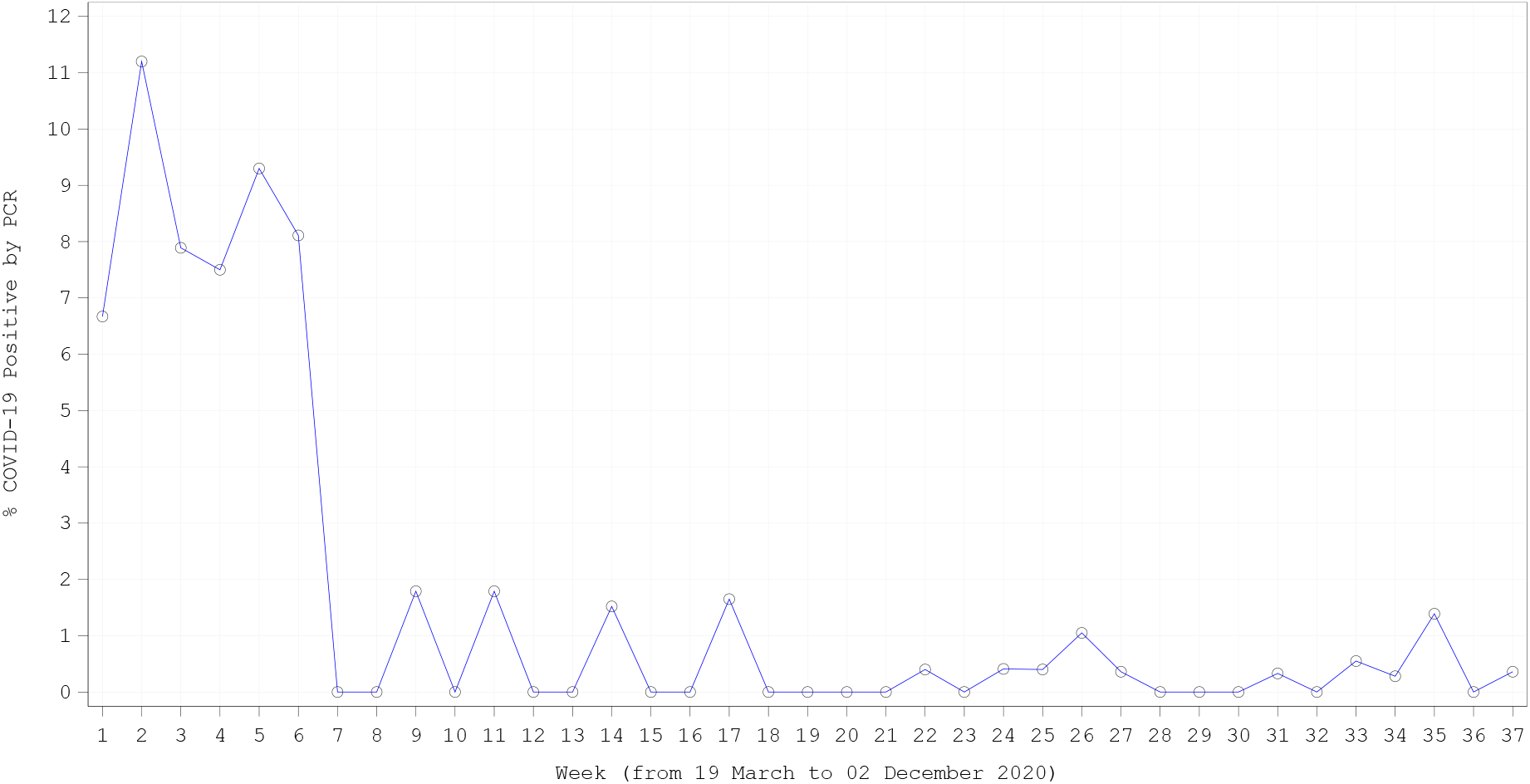
New positive SARS-CoV-2 tests measured by PCR.

Of the 6061 membrane antibody tests we conducted, 4953 (82%) were negative, 385 (6%) were IgG-positive only, 64 (1%) were IgM-positive, and 298 (5%) were positive for both IgG and IgM.

### Chemiluminescent Microparticle Immunoassay Results

Our data demonstrated an overall trend of waning IgG titres (**Figure 2**). After two months, titres were on average three times smaller than at their peak. The majority of volunteers showed a decline in antibody titres over the 90 days after symptom onset, and this remained low. We saw increasing titres in one individual and no change in titres for another two individuals. For five individuals, only one data point was available. Because of our small sample size, we applied a generalised estimating equation model [6] to estimate an antibody half-life of 70.4 days. Similar results have been recorded by other longitudinal studies, in which a peak in IgG titre was observed after 10-15 days [1,2,7,8], followed by a waning IgG response over the following two to three months [1,2].

**Figure 2.**
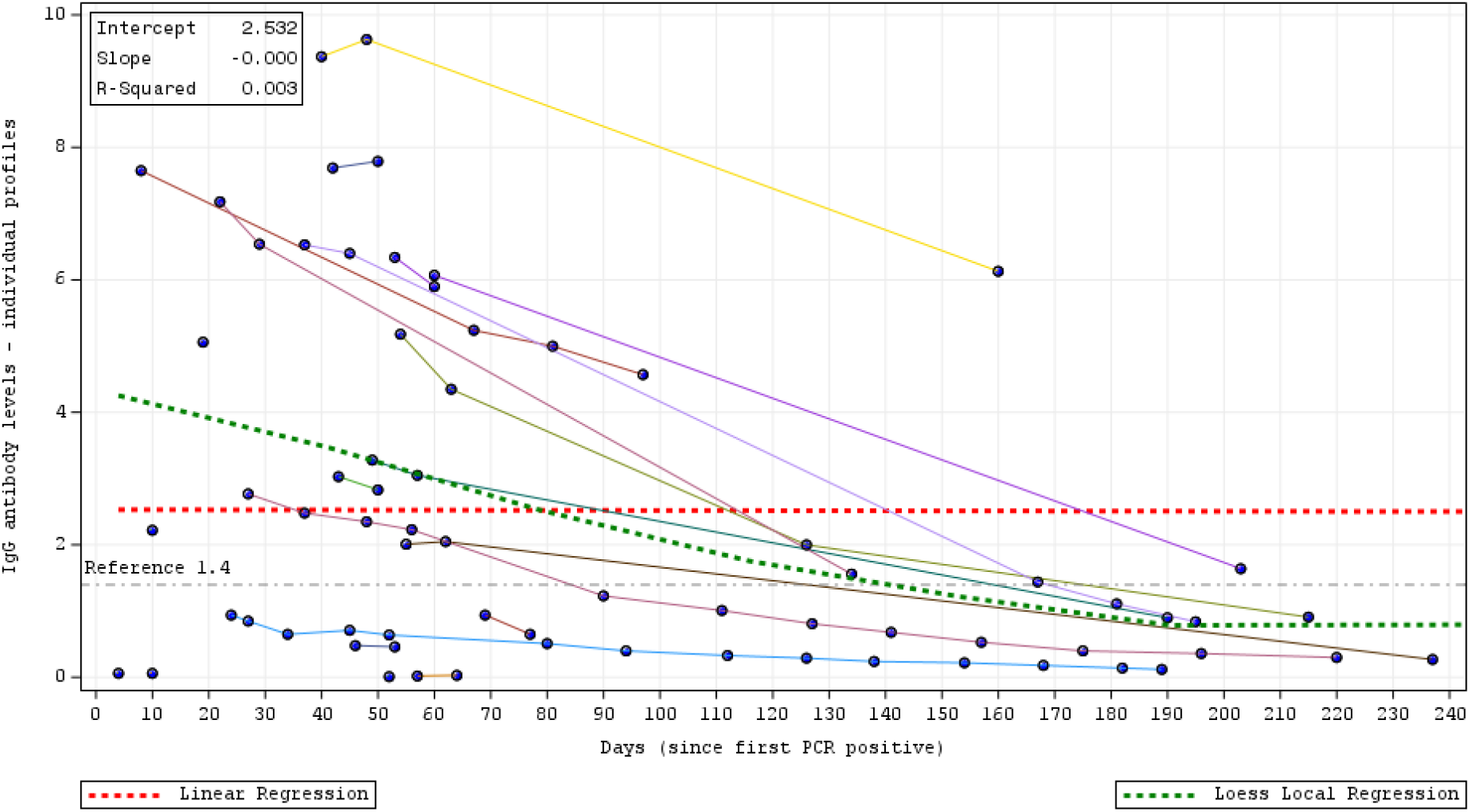
Individual IgG antibody levels (arbitrary units based on index value of Abbott COVID19 IgG Index S/CO) versus days since first PCR positive result. The fitted regression line reveals an overall trend of waning IgG titres for the 20 individuals measured.

Individuals confirmed positive for SARS-CoV-2 infection by PCR, membrane immunoassay, and via diagnosis by a physician according to their symptoms, had S/CO values below 1.4. Patients who were confirmed negative for SARS-CoV-2 by these methods had S/CO values 0.01 or less; the highest S/CO value for a negatively tested individual was 1.22. According to qualitative guidelines by Abbott, these patients would be declared negative for SARS-CoV-2 IgG.

## Discussion

We believe that our data makes a valuable contribution to gain a more rounded picture of the state of post-infection immunity to SARS-CoV-2 infection. What emerges is the need to be able to properly distinguish between post-COVID and other viral infections. Although studies have shown that the titre of circulating neutralising antibodies diminished after infection, it remains unclear whether the strength of the humoral response, as measured by antibody titre, correlates with favourable COVID-19 outcomes [9,10]. It seems likely that humoral immunity as mediated by circulating IgG should not be the only parameter measured when determining post-infection immunity. Nonetheless, waning antibody titres must be considered. Further studies of SARS-CoV-2 immunity are urgently needed to help guide public health policies and vaccine strategies in the future to minimise the spread of the viral infection. What remains to be uncovered is the level of antibodies required for a protective response. In vaccine trials, a positive significant correlation between anti–SARS-CoV-2 spike protein IgG measured by standardized ELISA and live virus SARS-CoV-2 microneutralisation assay was seen [11]. Should neutralising antibodies be shown to be protective in humans, routine serological assays (including Abbott’s CMIA) could be used for the standardised evaluation of functional antibodies by vaccine candidates in clinical trials.

By applying semi-quantitative tests that promise rapid results based on a fixed cut-off, such as the Abbott CMIA, clinicians and patients gain results that either classify them as negative or positive for COVID-19. Abbott’s SARS-CoV-2 IgG CMIA has shown 99.9% specificity and 100% sensitivity for detecting the IgG antibody in people 17 days after symptoms began [4] and a study comparing Abbott’s SARS-CoV-2 IgG CMIA using ELISA showed that the Abbott assay has a higher specificity (99.6%) than ELISA (91.5%) [12]. Our data also suggest that a number of patients who were confirmed to be COVID-19 positive appeared to be negative when using the Abbott CMIA test, suggesting that the fixed cut-off for a positive diagnosis has been set too high. The reason the threshold is too high could be that validation of the assay may have been conducted in patients with highly symptomatic COVID-19, rendering the existing threshold unsuitable for the wider population, which is a previously seen problem with SARS-CoV-2 antibody tests [13].

As the list of long-term effects of COVID-19 grows, it becomes vital to accurately determine if a patient was infected, because even asymptomatic or mild cases are associated with deleterious long-term effects in organs such as the lungs [10] and brain [14]. We do not necessarily suggest that any of these patients with low antibody titres have COVID-19 immunity – to the contrary, we advise all patients to continue to use all precautions advised. However, an accurate assessment of past SARS-CoV-2 infection is important to avoid false diagnosis of patients as SARS-CoV-2-negative. Uncertainty involving the reliability of antibody testing in the case of COVID-19 highlights the need to develop standardized diagnostic tests.

## Data Availability

Data available upon request.

